# A systematic review of the reporting and methodological quality of studies that use Mendelian randomisation in UK Biobank

**DOI:** 10.1101/2022.04.25.22274252

**Authors:** Mark J Gibson, Francesca Spiga, Amy Campbell, Jasmine N Khouja, Rebecca C Richmond, Marcus R Munafò

## Abstract

**Background:** Mendelian randomisation (MR) is a method of causal inference that uses genetic variation as an instrumental variable (IV) to account for confounding. While the number of MR articles published each year is rapidly rising (partly due to large cohort studies such as the UK Biobank making it easier to conduct MR), it is not currently known whether these studies are appropriately conducted and reported in enough detail for other researchers to accurately replicate and interpret them.

**Methods:** We conducted a systematic review of reporting and analysis quality of MR studies using only individual level data from the UK biobank to calculate a causal estimate. We reviewed 64 eligible articles on a 25-item checklist (based on the STROBE-MR reporting guidelines and the Guidelines for performing Mendelian Randomisation investigations). Information on article type and journal information was also extracted.

**Results:** Overall, the proportion of articles which reported complete information ranged from 2% to 100% across the different items. Palindromic variants, variant replication, missing data, associations between the IV and variables of exposure/outcome and bias introduced by two-sample methods used on a single sample were often not completely addressed (<11%).

There was no clear evidence that Journal Impact Factor, word limit/recommendation or year of publication predicted percentage of article completeness (for the eligible analyses) across items, but there was evidence that whether the MR analyses were primary, joint-primary or secondary analyses did predict completeness.

**Conclusions:** The results identify areas in which the reporting and conducting of MR studies needs to be improved and highlights that this is independent of Journal Impact Factor, year of publication or word limits/recommendations.

## Introduction

Mendelian randomisation (MR) is a method of causal inference that uses genetic variation as an instrumental variable (IV) for an exposure to estimate a causal effect. In principle (i.e., under certain assumptions), this estimate is free from confounding, including reverse causation (1). It is still a relatively new technique and, due to advances in genetic research, it is becoming more popular and widely used (2). Large cohort studies, such as the UK Biobank (UKB) which contains genetic, health and lifestyle data for around half a million people, have made it relatively easy to perform powerful MR analyses quickly (3). UKB’s aim is to encourage as many *bona fide* researchers as possible to use the resource for health-related research. As such, it provides the opportunity to access and analyse data which can be used to evaluate the reproducibility and robustness of published findings. While reporting guidelines such as the STROBE-MR (4) have recently been developed, it is not currently known whether MR studies report their analyses appropriately and report them in enough detail for others to accurately replicate and interpret them.

One previous systematic review found that only 44% of MR studies discussed the plausibility of the core assumptions and 14% gave insufficient detail of the statistical analysis (5). However, this review only looked at articles pre-2014, before both UKB became available and before MR became popular and widely used in epidemiology. Furthermore, while this review looked at a broad spread of MR methodologies, it only assessed the articles on these two points meaning it’s focus was rather narrow. Another review assessed the reporting quality of MR articles (up to 2017) on cancer outcomes and found around half the articles included (40% – 69%) did not report subject characteristics, did not conduct power calculations, did not describe the core MR assumptions and did not exclude variants that diverged from Hardy-Weinberg equilibrium (6). This review assessed more recent articles and assessed these articles in more detail but focused on the narrow topic of cancer MR studies.

The aim of our systematic review is to assess whether published articles that conduct MR analyses using individual-level data from the UKB cohort use appropriate analyses and report enough details to allow accurate interpretation and replication and whether this varies across article type. The findings will highlight which specific characteristics of MR analyses are omitted. As MR is a rapidly expanding field, it is vital we make sure the research is being accurately conducted and reported so it can be adequately interpreted and replicated. This will lead to work in the field being more robust, which in turn will lead to a reduction in wasted resources and an increase in impact.

## Methods

As this review does not investigate a particular exposure-health outcome relationship but instead investigates reporting and analysis quality of MR studies across fields, this review was not registered with a systematic review register, no synthesis was conducted, and no risk of bias assessment was completed. Where applicable we followed the 2020 PRISMA reporting guidelines.

### Article search and eligibility criteria

We searched four databases (Web of Science, EMBASE, PubMed and PsycINFO) for articles which contained “UK Biobank”, “UKB” or “UKBiobank” and “Mendelian randomisation” or “Mendelian randomization” on the 04/11/2020 (Supplementary Table S1).

We only included articles that conducted an MR analysis exclusively using individual-level UKB data to obtain a causal estimate of the exposure-outcome relationship. This includes articles which apply two-sample MR methods on UKB data either by utilising a split-sample approach or using the same UKB sample to estimate the IV-exposure and IV-outcome associations. We excluded:

- Articles where UKB data was only used when pooled with data from other cohorts.
- Articles where only an IV-outcome regression was conducted as the MR analysis.
- Articles where only two-sample MR was conducted using an external sample.
- Articles where the MR analysis was only conducted on summary-level data (i.e., not individual-level data).
- Articles where there were more than 10 independent exposures and/or outcomes (e.g., phenome-wide association studies).
- Any publication types (e.g., conference abstracts, meta-analyses, reviews, preprints, letters, editorials) that are not full research articles.
- Retracted articles.
- Articles not in English.
- Articles published before UKB was launched (30^th^ March 2012).
- Articles for which the full text or supplementary material cannot be accessed.

Article eligibility was assessed by one reviewer for title and abstract screening, and by two reviewers independently for full-text screening, with a third reviewer resolving any conflicts.

### Data extraction

Data extraction was carried out by two independent reviewers for each article with any conflicts being resolved by a third reviewer. Articles were reviewed using a 25-item checklist (Supplementary Table S2). On each item, the reviewer answered either “Yes”, “Partially” or “No”, with “Unclear” or “NA” also being allowed responses for specific items.

Both the STROBE-MR guidelines (4) and the “Guidelines for performing Mendelian Randomisation investigations” (7) were used to create the data extraction items. Each item is based on an item or items from the STROBE-MR while the “Guidelines for performing Mendelian Randomisation investigations” were used to finalise the wording of each question to make sure it covers appropriate analytical practices as well as reporting practices. To measure how reporting quality differs across journals and article types, the Journal Impact Factor, open access status of the article and journal word limit/recommendation were extracted by the primary reviewer and whether the authors claimed to follow STROBE-MR guidelines, and whether the analysis was a primary, joint primary or follow-up analysis was also dual extracted.

First, a project protocol including initial data-extraction items was created and pre-registered on the Open Science Framework (https://osf.io/hzfj7/). Then a pilot of ten articles was conducted to finalise the data-extraction items. During data-extraction the wording of some items was altered slightly to remove ambiguity in certain cases, while maintaining the intended meaning. We extracted information on:

- The reporting of the three core MR assumptions.
- Whether a one or two-sample design was used.
- The variables of interest (the exposure-outcome relationship, the UKB variables used, and how these variables were handled).
- The sample (exclusions, size, and genetic quality control).
- The instrumental variable (variants/weights used, selection of variants, replication of variants, proxy variants, and palindromic variants).
- The analysis method (MR estimator and covariates).
- How bias was addressed (missing data, multiple testing, variant heterogeneity, instrument strength, sensitivity analysis, and sample overlap).
- The reporting of results (descriptive statistics, instrument-exposure and instrument-outcome associations, the causal effect estimate, measures of uncertainty, the units, and figures).
- The reporting of software and code (including versions used).

### Statistical analysis

The percentage of articles which obtained a “Yes”, “Partially” and “No” (also “Unclear” where relevant) on each item of interest was calculated. Linear regression was then used to investigate potential associations between the completeness of articles (average percentage across items for each article with “Yes” as 100%, “Partially” as 50% and “No” or “Unclear” as 0%) and the 2020 Journal Impact Factor (logged), word limit/recommendation (both number of words and whether a word limit/recommendation was present or not) and whether the MR analysis was the primary, joint primary or secondary analysis. One paper published in a journal which had a limit of 35 pages was classed as having no word limit/recommendation. Due to the lack of variance within open access status (“Yes” = 81% [52], “Partially” = 12% [8] and “No” = 6% [4]) and whether the authors followed the relevant reporting guidelines such as the STROBE-MR (“Yes” = 2% [1], “Partially” = 8% [5] and “No” = 91% [58]) regressions were not run using these as predictors. For the latter, “Partially” refers to use of general reporting checklists not specific to MR. All analyses were carried out in R version 4.1.0 and are available along with the data at https://codeocean.com/capsule/1654662/tree/v1.

### Results

A total of 64 articles were included in the final sample (see figure one for a flow chart of article exclusion and Supplementary Table S3 for a list of articles excluded at full-text screening or after with reasons for exclusion). The articles included in the review and the data extracted for each article can be seen in Supplementary Table S4. Overall, the mean article completeness was 56% (standard deviation = 9%). Across second reviewers, mean completeness was consistent 54% to 58% (SD = 9% to 10%). Percentages for each item can be seen in Supplementary Table S5 and Figure 2.

**Figure 1.**
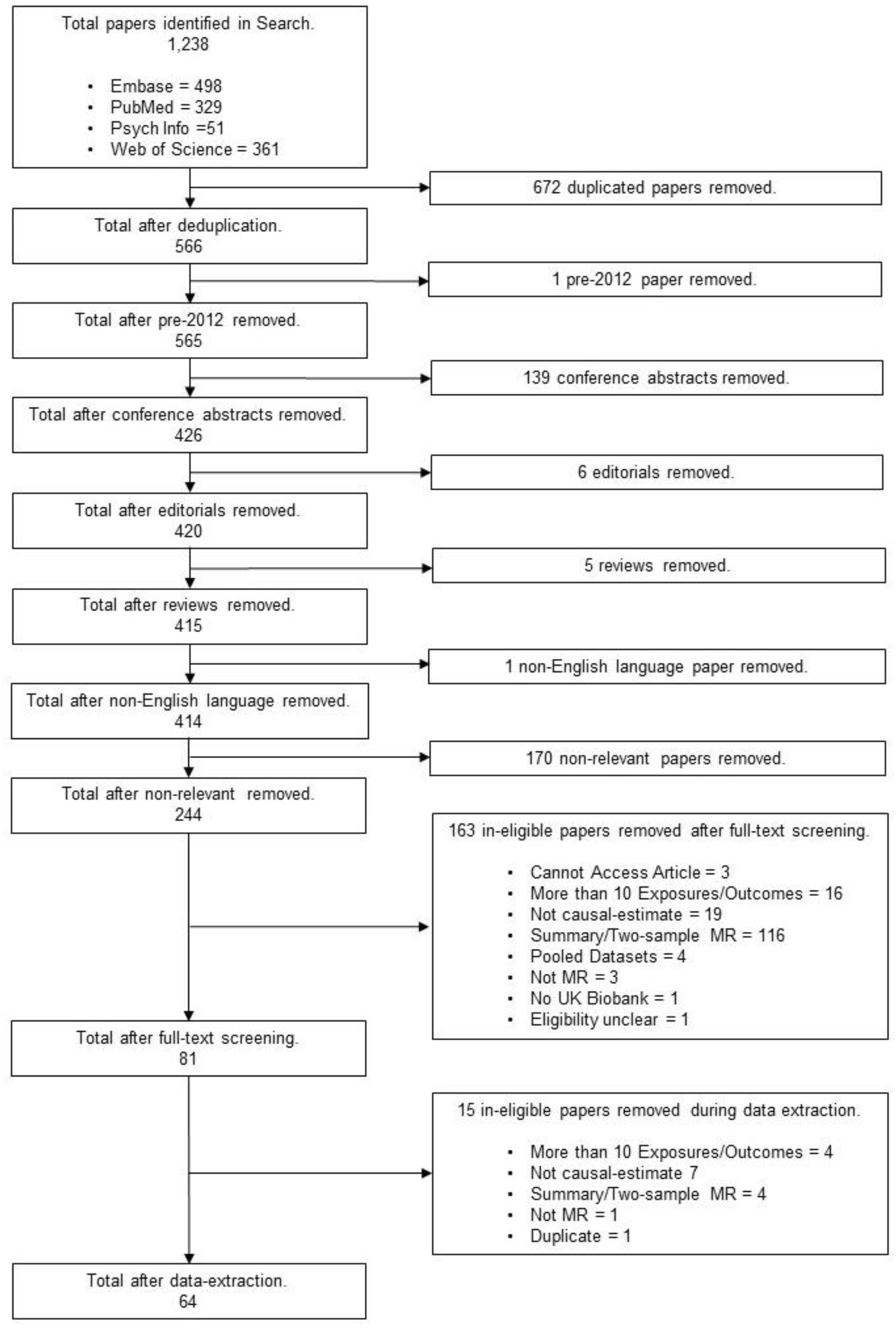
Flow-chart of screening.

**Figure 2.**
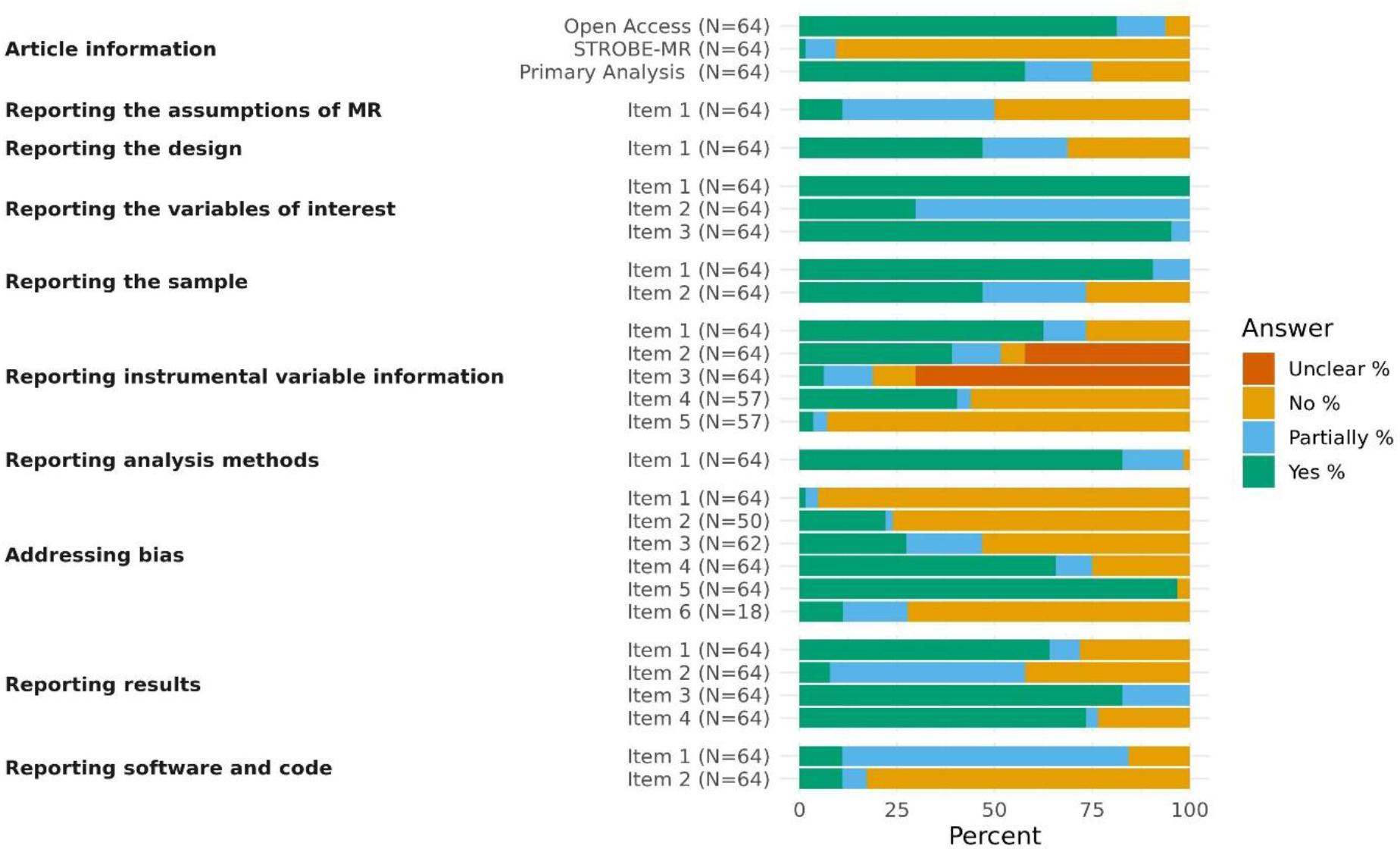
Percentages of each response for each data extraction item. *Note: The number of articles for each item varies due to some items not being applicable for all articles*.

### Reporting the assumptions of MR

Of the 64 articles included in this review, only 11% (7) of articles completely reported the three core assumptions of MR, while 39% (25) partially reported them (i.e., reported them incorrectly) and 50% (32) did not outline all three assumptions / did not outline them at all. While these assumptions have been previously detailed in a number of articles and reporting them it is not necessary for replication, they are important for the interpretation of the results and will not be common knowledge to those who do not conduct MR themselves. Therefore, it is still good practice to report them. Often however, they are reported wrong which arguably does more harm than good. The second assumption is regularly reported as “no association between the IV and confounders of the exposure-outcome relationship”, instead of the correct assumption, “there are no confounders of the IV-outcome association”. The distinction between these assumptions is that the former is a specific form of pleiotropy that is already covered by the third assumption, whereas the latter covers important and otherwise unmentioned issues such as population stratification and dynastic effects (8).

### Reporting the design

Articles regularly did not clearly report whether the study was a one-sample or two-sample MR design. While 47% (30) did report this accurately, 22% (14) only partially reported this (i.e., implied its design in reference to it not being the alternative) and 31% (20) did not. This information is not only trivial to provide but helps to clearly communicate how the analysis was conducted (and therefore how it can be replicated) without having to infer this from more complicated details. It is also important to understanding the biases the results may be subject to and, thus, it is vital for the interpretation of the results. Therefore, reporting this should be a basic requirement of MR research.

### Reporting the variables of interest

As would be expected, 100% of articles completely reported what exposure outcome relationship was being assessed. However, only 30% (19) of articles completely reported which UKB phonotypes were used versus 70% (45) which partially reported this (i.e., did not provide the UKB field IDs). UKB has a number of similar and closely related variables and without the IDs it is difficult to be certain which exact variable has been used by researchers. Reporting of field IDs is the only way to completely remove this ambiguity and should be seen as a necessity when using UKB data; 95% (61) of articles clearly reported how these variables were handled in enough detail to replicate the analysis and interpret the results, versus 5% (3) which only partially reported this.

### Reporting the sample

For information on the eligibility criteria and subsample size, 91% (58) of articles completely reported this information while 9% (6) only partially reported this. As sample size and exclusion criteria are vital to reporting in all fields of science this high rate is unsurprising. In contrast, only 47% (30) of articles completely reported information on the genetic data (i.e., exclusion of variants and imputation information) while 27% (17) partially reported this information and 27% (17) didn’t report it at all. As these processes in most cases were conducted by UKB centrally, most researchers may feel this will not affect replication attempts and there is little need to report this. However, if what has been done is not reported, it is unclear if the researchers have done extra processing to the data and it hinders interpretation of the results for those unfamiliar with UKB. Therefore, it is good practice to report this (albeit briefly).

### Reporting instrumental variable information

Of the 64 articles included in the study, 62% (40) completely reported the genetic variants and weights used to construct the IV, while 11% (7) partially reported this and 27% (17) did not. 39% (25) reported that variants were appropriately selected and weighted (i.e., identified in a different sample as that used in the analysis, or externally weighted), 12% (8) reported only partially appropriate selection (i.e., identified in the same sample and unweighted or identified and weighted in a larger sample which includes the sample used in the MR analysis), 6% (4) did not appropriately select variants (i.e., identified and weighted these in the same sample used in the MR analysis) and 42% (27) did not give enough detail to assess this. Whether the variants used had been independently replicated had lower reporting quality across articles; 6% (4) of articles reported that variants were independently replicated, while 12% (8) reported they were partially replicated (e.g., replicated in a partially overlapping sample), 11% (7) used un-replicated variants and 70% (45) did not report this.

Variants which were identified in the sample used for the analysis, or which were not replicated, should be only used if there is no other option. If unavoidable, this should be made clear, and whether this has introduced bias into the results should also be addressed. For the 57 articles for which information on proxies and palindromic variants were relevant (i.e., used variants for the IV which were not identified in UKB), 40% (24) of articles clearly reported proxy information (i.e., whether all variants were present in UKB and, if not, whether these variants were excluded or proxied), 4% (2) partially reported this and 56% (32) did not. For the reporting of palindromic variants, 4% (2) clearly reported this if they were present and how they were handled if so, 4% (2) partially reported this and 93% (53) did not.

Reporting all of this information should be considered standard practice, however, this was clearly not the case. A reason for the low rates of reporting details on proxy and palindromic variants may be that researchers feel that if these were not present they do not need to be mentioned, and therefore to omit this information is as good as saying they were not an issue for the study. This is poor practice as it creates ambiguity, especially when a large number of variants are being used.

### Reporting analysis methods

Of the 64 articles, 83% (53) clearly reported the MR estimator used and the covariates adjusted for, while 16% (10) partially reported this and 2% (1) did not. While this rate of reporting is relatively high compared to the other items included in this review, this is fairly low considering how vital to replication and interpretation this information is.

### Addressing bias

Whether missing data could have biased the results was clearly addressed by 2% (1) of articles, while 3% (2) partially addressed this and 95% (61) did not (these articles may still have presented the percentages of missing data but did not comment on the impact of this issue). Of the 50 articles which conducted multiple testing, 22% (11) addressed this (i.e., corrected for this bias or explained why no correction was needed), 2% (1) partially addressed this and 76% (38) did not. For the 62 articles for which it was applicable (i.e., more than one variant was used), 27% (17) addressed heterogeneity (i.e., reported heterogeneity or used a method which excluded outliers) of the individual variant MR estimates, while 19% (12) partially did and 53% (33) did not. Reporting of assessments of instrument strength (i.e., the F-statistic or the R^2^/variance explained) was better, with 66% (42) of articles completely reporting this, 9% (6) partially reporting this and 25% (16) not reporting this. Furthermore, the conducting and reporting of sensitivity analysis was common across articles, with 97% (62) completely reporting this and 3% (2) not. However, sensitivity analysis is a broad category of analyses, and this does not mean the analyses were conducted well or were the best suited sensitivity analyses for the study in question. The quality of sensitivity analyses used was not assessed, and this is of a large enough scope to be the focus of an entire systematic review. Finally, of the 18 articles which used two-sample MR methods on one-sample data, only 11% (2) of articles addressed this potential source of bias, while 17% (3) partially addressed this and 72% (13) did not.

### Reporting results

Descriptive statistics were completely reported by 64% (41), partially reported by 8% (5) and not reported by 28% (18) of articles. Reporting rates were far worse for the IV-exposure and IV-outcome associations which were completely reported by 8% (5), partially reported (i.e., either only one of the two reported or reported for each individual variant) by 50% (32) and not reported by 42% (27) of articles. For reporting of the causal estimate reporting rates were (as would be expected) far better, with 83% (53) of articles completely reporting this and 17% (11) partially reporting this (i.e., not making the units clear, not reporting an interpretable scale, or not giving a measure of uncertainty). Finally, 73% (47) of articles visualised the results in a figure, with 3% (2) only partially doing this (i.e., the figure was not the best choice for visualisation or was difficult to interpret) and 23% (15) not visualising the results at all. While visualising the results is not vital to replication, it does aid interpretation and researchers should be doing all they can to help communicate their findings as clearly as possible.

### Providing software and code

Of the articles, 11% (7) clearly reported the statistical programs, packages and versions of each used for the statistical analysis, 73% (47) partially reported this (missing versions or packages) and 16% (10) did not. If complete code is provided then omissions of packages used and version numbers would matter little, however, only 11% (7) of articles provided complete code, 6% (4) provided partial code and 83% (53) did not provide any code. The provision of complete code is a very simple solution to the issue of replicability and would cover most of the other items in this review. Moving forward, this is the main component of open science which is needed to improve replicability in the field.

### Effects of article Type and Journal

There was no clear evidence that 2020 Journal Impact Factor, word limit/recommendation (both word number or whether a limit/recommendation was present) or year of publication predicted percentage of article completeness across items, but there was evidence that primary analysis status (“Yes” = 58% [37], “Partially” = 17% [11] and “No” = 25% [16]) did increase completeness (mean difference in completeness percentage [95% confidence interval] = 7% [2%, 12%]). For primary analysis status, “Partially” refers to the MR analysis being a joint primary analysis and was coded as 0.5. While it might be expected that articles which did an eligible MR analysis as a joint primary analysis or secondary analysis would report this analysis less completely than those which conducted MR as the sole primary analysis, the ability to report methods and results in supplementary materials means there is no excuse for not reporting analysis and results fully. This finding implies that researchers do not make proper use of supplementary materials and do not report all they should in these. Regression results can be seen in Supplementary Table S6 and Figure 3.

**Figure 3.**
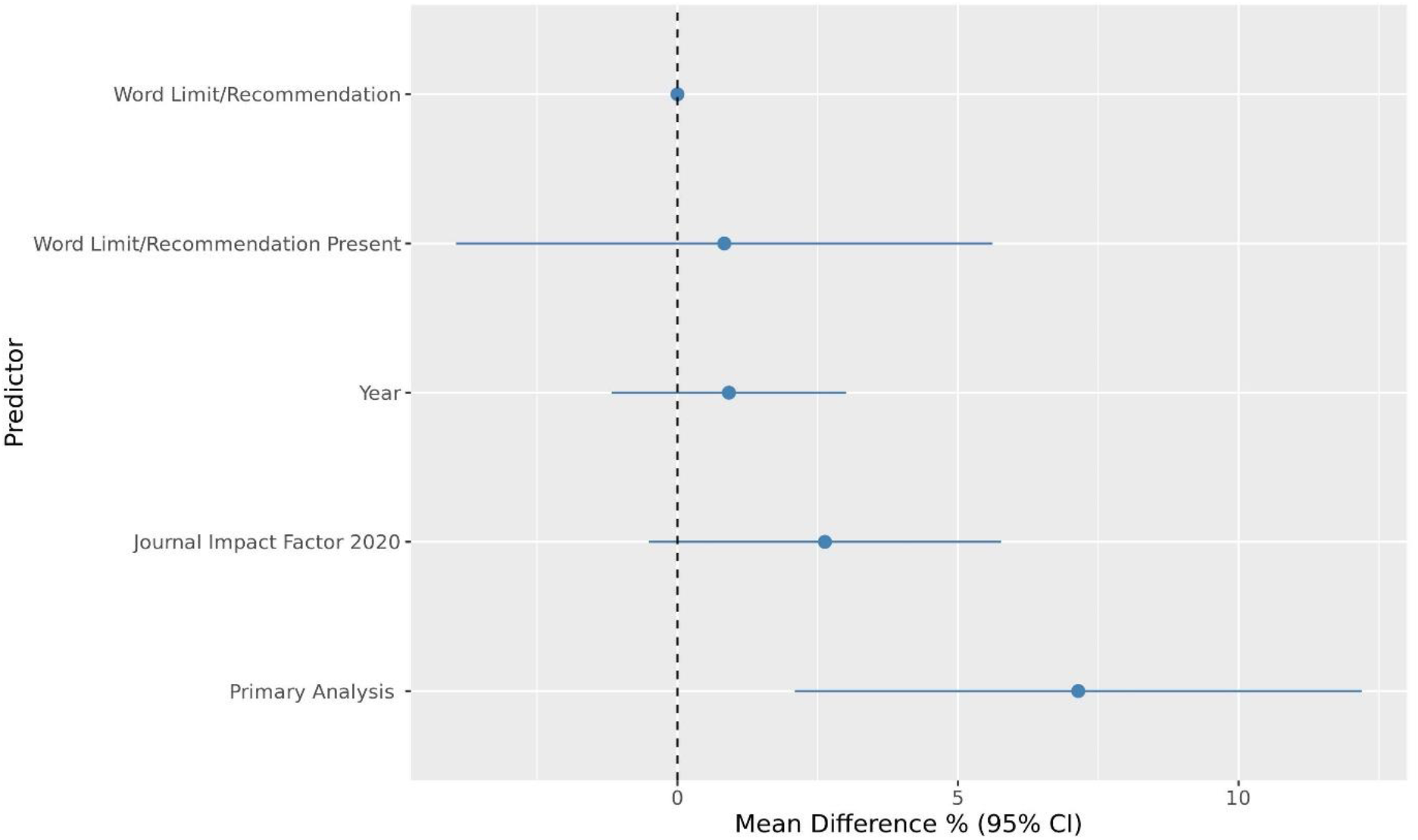
Association of predictors and article completeness. *Note: N = 64 for all except word limit/recommendation where N = 42; 2020 Journal Impact Factor logged due to skewness*.

## Discussion

This study shows that the quality of analyses and reporting was variable across items and was worst for aspects relating to palindromic variants, variant replication, missing data, associations between the IV and variables of exposure/outcome and bias introduced by two-sample methods used on a single sample. Article completeness was not predicted by Journal Impact Factor, word limit/recommendation or year of publication, but was predicted by primary analysis status. The study highlights areas which need to be improved going forward to improve the quality of research in the field.

The results of this systematic review roughly align with those of previous, less comprehensive reviews. A previous study found that 44% of MR articles discussed the plausibility of the core assumptions (5), while our study did not look at this specifically, we found that 50% outlined the core assumptions (albeit mostly incorrectly), 27% completely addressed variant heterogeneity (relevant to the third core assumption), and 66% completely addressed the strength of the IV (i.e., the first assumption). However, the previous study did not investigate the reporting of variant heterogeneity like we did, and also found lower reporting of instrument strength than us (30-34%). The previous study also identified that 14% of studies gave insufficient detail of the statistical analysis (i.e., information to accurately replicate including the confidence intervals of the results), while we found that 18% did not give complete information about the MR estimator and covariates and 17% did not provide the causal estimate on an interpretable scale with measures of uncertainty. Another review (on oncology MR studies) found that 49% of articles reported subject characteristics and 48% did not describe the core MR assumptions. We found similar but higher rates of reporting of subject characteristics (64%) and almost identical rates of reporting the core assumptions (50%) in a broader sample of MR papers. It is important to note that no papers from the current review were included in the previous reviews.

A strength of this study is that data were dual-extracted, and conflicts were resolved by a third reviewer, reducing the impact of bias and human error. The study is also strengthened by the comprehensiveness of the review items. However, while this review includes a large number of MR papers (64) across all fields, it focuses on a very specific subset: those which used individual-level UKB data only to obtain a causal estimate for less than 10 exposures or outcomes. To include papers outside of this subset would have made the review too large but it is possible that article completeness differs drastically outside this subset.

Furthermore, several items could have been coded in more detail if the scope of the project was not already so large (i.e., the quality of sensitivity analysis or code provided could have been extracted). The replicability of articles would also be better assessed by carrying out full replication attempts. These are two areas which should be followed up in future research, if only on a small subsample of articles. It would also be worthwhile to investigate whether the creation of the STROBE-MR improves paper completeness, however, a greater period of time needs to pass from its creation for this to be feasibly investigated.

To conclude, the findings of this study highlight areas of poor conduct and reporting in MR research which need to be improved to increase replicability and impact. Furthermore, these appear to be consistent across Journal Impact Factor and word limit/recommendations as well as the year of publication. Only primary analysis status predicted article completeness implying that researchers do not sufficiently use supplementary materials to report secondary analysis and results. Increased analysis and reporting quality in the field is vital for improving our ability to replicate and accurately interpret findings, increasing the impact of research and making better use of public money. Future research should focus on the quality of certain aspects such as code or sensitivity analysis, as well as attempting direct replications, and should investigate the impact of the STROBE-MR specifically.

## Supporting information

2020 PRISMA reporting guidelines

## Data Availability

The data is available via the University of Bristol online data repository and on https://codeocean.com/capsule/1654662/tree/v1 as well as in Supplementary Table S4.

## Footnotes

### Contributor and guarantor information

MJG is the primary investigator and drafted the protocols, while FS, RCR and MRM commented on and edited these. MJG caried out title/abstract screening. MJG, FS and RCR carried out full-text screening and the data-extraction pilot. Final data-extraction was carried out by MJG, FS, AC and RCR. The analysis was then carried out and the manuscript was written by MJG with guidance from JNK, RCR and MRM. The corresponding author attests that all listed authors meet authorship criteria and that no others meeting the criteria have been omitted.

